# COVID-19 infections following outdoor mass gatherings in low incidence areas: retrospective cohort study

**DOI:** 10.1101/2020.10.22.20184630

**Authors:** Oren Miron, Kun-Hsing Yu, Rachel Wilf-Miron, Nadav Davidovitch

## Abstract

**Objective:** Indoor mass gatherings in counties with high COVID-19 incidence have been linked to infections. We examined if outdoor mass gatherings in counties with low COVID-19 incidence are also followed by infections.

**Methods:** We retrospectively examined COVID-19 incidence in 20 counties that held mass gathering rallies (19 outdoor and 1 indoor) in the United States in August-September 2020. They were compared to the rest of the United States counties. We utilized a 7-day moving average and compared the change on the gathering date and 15 days later, based on the 95% confidence interval. For control counties we used the median of the gathering dates.

**Setting:** The United States

**Population:** 8.4 million in the counties holding mass gatherings, and 324 Million in the rest of the counties in the United States.

**Main Outcome Measure:** Change in COVID-19 incidence rate per 100,000 capita during the two weeks following mass gatherings.

**Results:** In the two weeks following the gatherings, the COVID-19 incidence increased significantly in 14 of 20 counties. The county with the highest incidence increase (3.8-fold) had the 2^nd^ lowest incidence before the gathering. The county with the highest decrease (0.4-fold) had the 3^rd^ highest incidence before the gathering. At the gathering date, the average incidence of counties with gatherings was lower than the rest of the United States, and after the gathering, it increased 1.5-fold, while the rest of the United States increased 1.02-fold.

**Conclusion:** These results suggest that even outdoor gatherings in areas with low COVID-19 incidence are followed by increased infections, and that further precautions should be taken at such gatherings.

**What is already known on the topic:** Mass gatherings have been linked to COVID-19 infections, but it is less clear how much it happens outdoors, and in areas with low incidence.

**What this study adds:** COVID-19 infections increased significantly in 14 of 20 counties that held mass gathering rallies in the United States, 19 of which were outdoors. The county with the highest incidence increase (3.8-fold) was outdoors and had a low incidence before the gathering. The average incidence of all 20 counties with gatherings was lower at the gathering day compared with the rest of the United State, and it increased 1.5-fold following the gatherings. Our findings suggest a need for precautions in mass gatherings, even when outdoors and in areas with a low incidence of COVID-19.

## Introduction

The United States limited most mass gatherings since March 2020 to prevent the spread of coronavirus disease 2019 (COVID-19).^1,2^ There have been mass gatherings in rallies that were linked to COVID-19 cases, but these gatherings were indoor and in counties with high COVID-19 incidence.^3^

It is unclear if outdoor mass gatherings in counties with low COVID-19 are followed by increased COVID-19 incidence. It is also unclear if the mass gathering effects seen earlier in the pandemic would be reduced by the effect of increasing precautions, such as social distancing and facemasks.^4,5^ This raises the need to examine the COVID-19 incidence following mass gathering in late 2020.

## Methods

We retrospectively examined daily COVID-19 incidence from every U.S. county, using the open data repository of Johns Hopkins University.^6^ We searched for counties with mass gatherings for rallies since August 2020 with at least 15 days of incidence data following the gatherings, since most infections are detected in this period.^7^ We found 20 counties meeting those criteria that held such mass gatherings from August 17 to September 30. Those counties had a combined population of 8,408,016 based on U.S. Census data. We analyzed the county incidence rate per 100,000 capita and utilized a weekly moving average to account for the change in days of the week. In each county, we examined if the incidence changed in the 15 days following the gathering date based on the 95% confidence interval. Lastly, we averaged the rate in all the counties that held these mass gatherings and compared it to all other U.S. counties. For non-gathering counties, we used a control-gathering date that was the median of the actual gathering dates (September 15, 2020).

## Results

In the 15 days following the gatherings, the incidence increased significantly in 14 of 20 counties (Table 1; Figure 1). The increase was 3.8-fold in Lackawanna, 3.7-fold in Beltrami, 3-fold in Marathon, 2.9-fold in Blue Earth, 2.1-fold in Winnebago, 2.1-fold in Dauphin, 2-fold in York, 1.9-fold in Rockingham, 1.9-fold in Montgomery, 1.6-fold in Clark (only indoor gathering), 1.6-fold in Allegheny, 1.4-fold in St. Louis, 1.3-fold in Westmoreland, and 1.2-fold in Duval.

**Table 1:** Study characteristics

| Gathering County | Gathering Site | Gathering date | County Population | Cases/100k day-0 | Cases/100k day-15 | Day 0-15 Change | p value |
| --- | --- | --- | --- | --- | --- | --- | --- |
| Lackawanna, Pennsylvania | Outdoor | Aug 20 | 209,674 | 2.2 | 8.2 | 3.8-fold | <0.001 |
| Beltrami, Minnesota | Outdoor | Sep 18 | 47,188 | 7.9 | 29.4 | 3.7-fold | <0.001 |
| Marathon, Wisconsin | Outdoor | Sep 17 | 135,692 | 17.4 | 51.6 | 3-fold | <0.001 |
| Blue Earth, Minnesota | Outdoor | Aug 17 | 67,653 | 15.2 | 44.3 | 2.9-fold | <0.001 |
| Winnebago, Wisconsin | Outdoor | Aug 17 | 171,907 | 7.1 | 15.3 | 2.1-fold | <0.001 |
| Dauphin, Pennsylvania | Outdoor | Sep 26 | 278,299 | 6 | 12.5 | 2.1-fold | <0.001 |
| York, Virginia | Outdoor | Sep 25 | 68,280 | 2.9 | 5.9 | 2-fold | 0.008 |
| Rockingham, New Hampshire | Outdoor | Aug 28 | 309,769 | 1.8 | 3.4 | 1.9-fold | <0.001 |
| Montgomery, Ohio | Outdoor | Sep 21 | 531,687 | 8.9 | 17.3 | 1.9-fold | <0.001 |
| Clark, Nevada | Indoor | Sep 13 | 2,266,715 | 9 | 14.6 | 1.6-fold | <0.001 |
| Allegheny, Pennsylvania | Outdoor | Sep 22 | 1,216,045 | 4.9 | 7.7 | 1.6-fold | <0.001 |
| St. Louis, Minnesota | Outdoor | Sep 30 | 199,070 | 19.7 | 28.3 | 1.4-fold | <0.001 |
| Westmoreland, Pennsylvania | Outdoor | Sep 03 | 348,899 | 3.2 | 4.1 | 1.3-fold | 0.039 |
| Duval, Florida | Outdoor | Sep 24 | 957,755 | 12.3 | 14.5 | 1.2-fold | <0.001 |
| Lucas, Ohio | Outdoor | Sep 21 | 428,348 | 7.1 | 7.8 | 1.1-fold | 0.159 |
| Cumberland, North Carolina | Outdoor | Sep 19 | 335,509 | 14.3 | 14.6 | 1-fold | 0.811 |
| Douglas, Nevada | Outdoor | Sep 12 | 48,905 | 3.8 | 3.5 | 0.9-fold | 0.774 |
| Forsyth, North Carolina | Outdoor | Sep 08 | 382,295 | 8.3 | 7.5 | 0.9-fold | 0.161 |
| Saginaw, Michigan | Outdoor | Sep 10 | 190,539 | 8.9 | 7.1 | 0.8-fold | 0.015 |
| Yuma, Arizona | Outdoor | Aug 18 | 213,787 | 16.2 | 6.5 | 0.4-fold | <0.001 |
| **United States Counties with gathering** | **19 of 20 Outdoor** | **Median Sep 15** | **8,408,016** | **8.6** | **12.8** | **1.5-fold** | **<0.001** |
| **United States Counties without gathering** | **None** | **Control Sep 15** | **324,457,671** | **12.9** | **13.1** | **1.02-fold** | **<0.001** |

**Figure 1:**
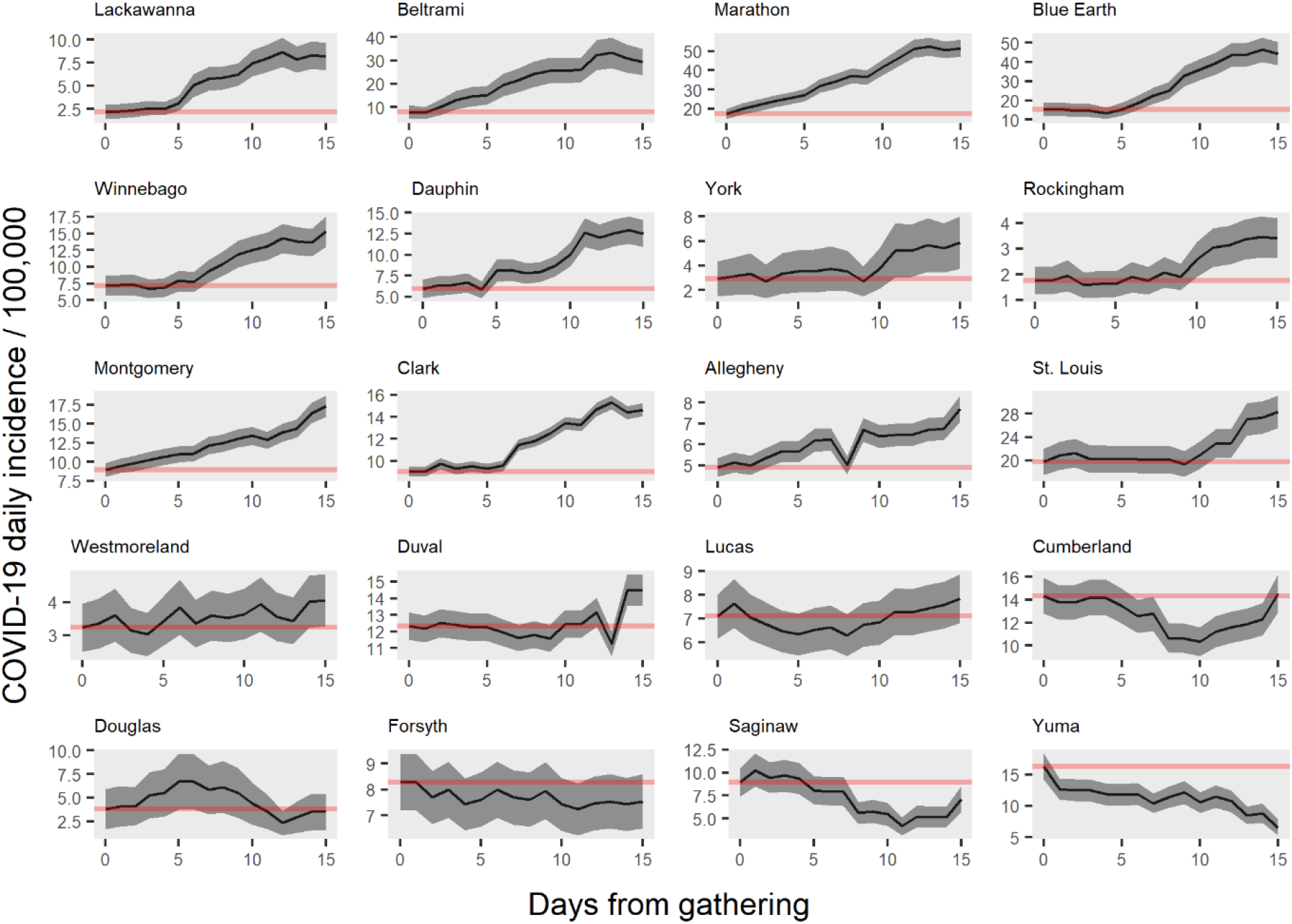
COVID-19 incidence rate following gathering by county. Legend: Coronavirus Disease-19 (COVID-19) incidence by days from gathering. Y-axis indicates COVID-19 daily incidence per 100,000 capita after a 7-day moving average. X-axis indicates the days from the gathering date. The red horizontal line indicates the incidence rate at the gathering date. The grey area indicates the 95%-confidence interval. The title above graph indicates the county shown.

In the 15 days following the gatherings, the incidence significantly decreased in 2 of 20 counties. The decrease was to 0.8-fold in Saginaw, and to 0.4-fold in Yuma.

The county with the highest relative increase (Lackawanna) had the 2^nd^ lowest an incidence at the gathering date, while the county with the highest relative decrease had the 3^rd^ highest incidence at gathering date (Yuma, Figure 2).

**Figure 2:**
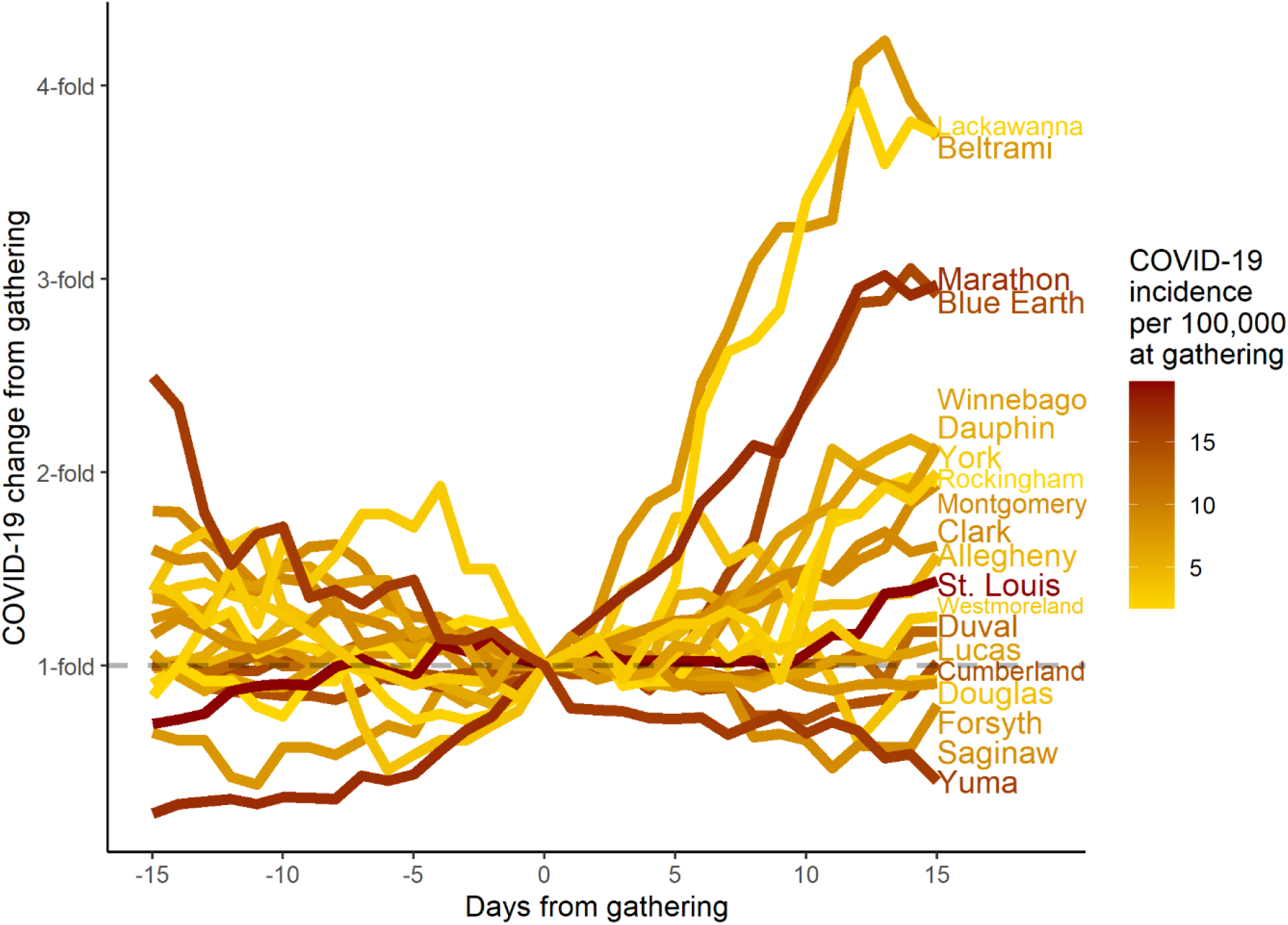
COVID-19 incidence change following gathering by county. Legend: Coronavirus Disease-19 (COVID-19) incidence by days from gathering. Y-axis indicates COVID-19 daily change from gathering date after a 7-day moving average. X-axis indicates the days from the gathering date. Dark brown indicates a higher COVID-19 rate at the gathering date, and yellow indicates a lower rate.

The average COVID-19 incidence of gathering counties at the gathering date was 8.6/100,000 capita, and 15 days after the gathering the incidence increased to 12.8/100,000 capita (1.5-fold increase). The incidence of the other U.S. counties at the control index date (September 15) was 12.9/100,000 capita, and 15 days later it was 13.1/100,000 (1.02-fold increase; Figure 3; Figure 4).

**Figure 3:**
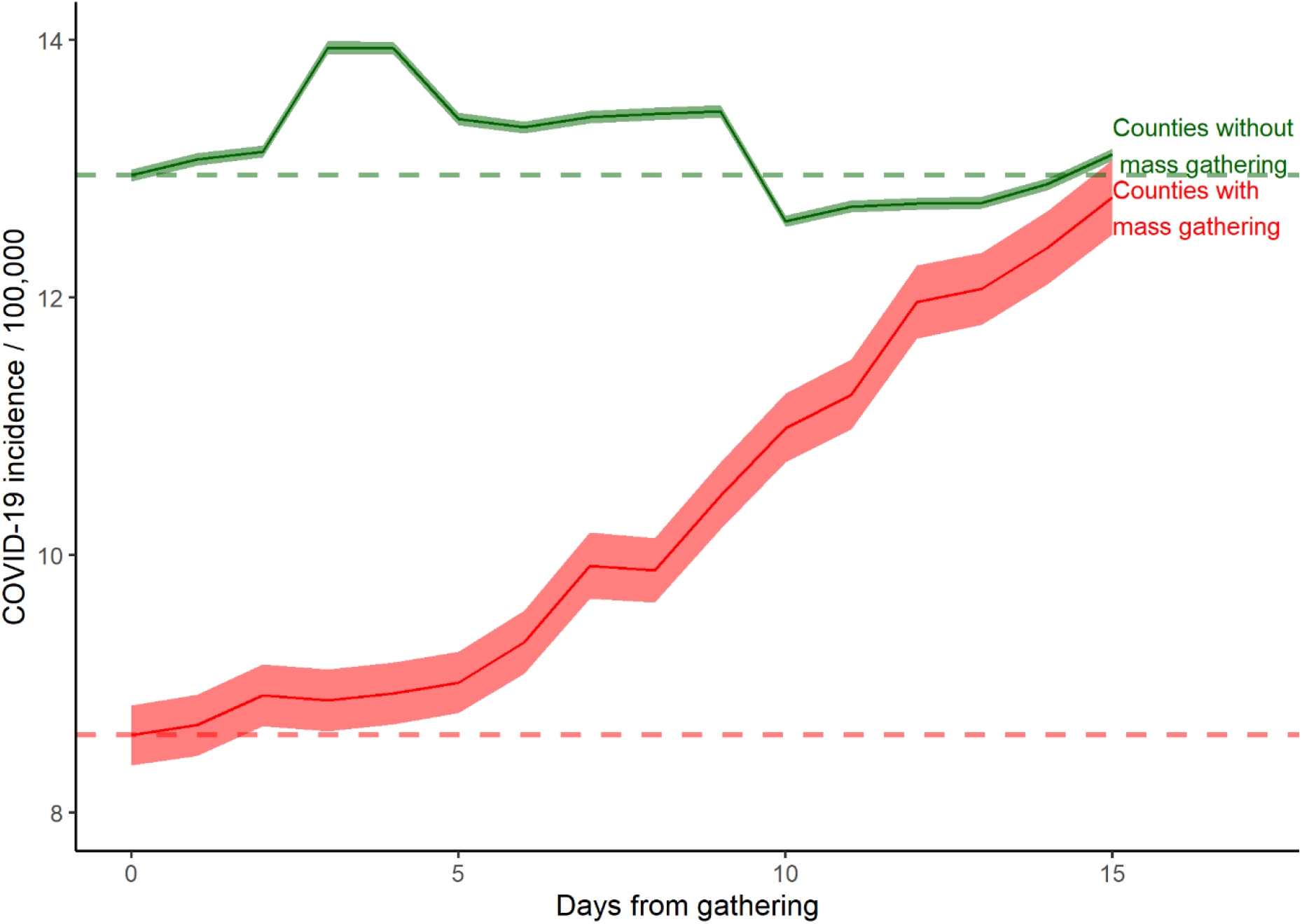
COVID-19 incidence rate in gathering counties and U.S. Legend: Y-axis indicates COVID-19 daily incidence per 100,000 capita after a 7-day moving average. X-axis indicates the days from the gathering date, with the counties without gatherings having a control index date of September 15 (the median of the gathering dates). Counties with the gatherings are in red, and counties without the gatherings are in green. The horizontal line indicates the incidence rate on the gathering date of each group.

**Figure 4:**
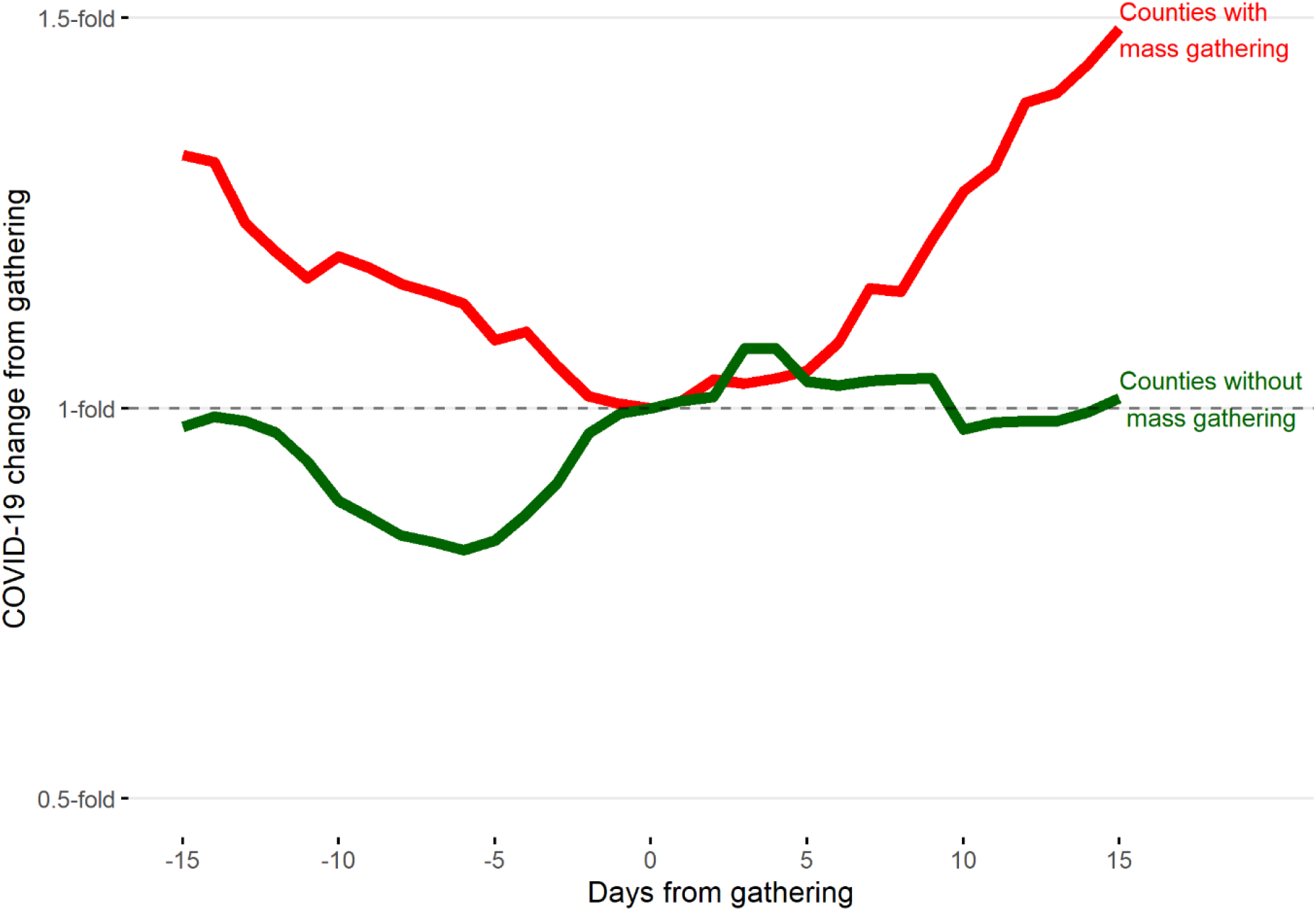
COVID-19 incidence change in gathering counties and U.S. Legend: Y-axis indicates COVID-19 daily incidence change from gathering date, after a 7-day moving average. X-axis indicates the days from the gathering date, with the counties without the gatherings having a control index date of September 15 (the median of the gathering dates). Counties with the gatherings are in red, and counties without the gatherings are in green.

## Discussion

Our analysis showed that 14 of 20 mass gatherings were followed by a significant increase in the COVID-19 incidence rate. The county with an indoor gathering had a significant increase, but the relative increase was higher in 9 outdoor counties. This suggests that placing a mass gathering outdoors does not entirely prevent the spread of COVID-19, and other precautions are warranted.

A common precaution to avoid COVID-19 infections is to hold gatherings in counties with low incidence.^2^ We found that the county with the 2^nd^ lowest COVID-19 incidence at the gathering day had the highest incidence increase after the gathering. Additionally, the county with the 2^nd^ highest incidence before the gathering had the largest decrease in incidence following the gathering. Taken together, this suggests that holding gatherings in low incidence counties is not always safe, especially if some attendees come from neighboring areas with higher incidence.^3,8^

The average incidence in gathering counties had a 1.5-fold increase after the gathering, while the incidence in the rest of the U.S. increased 1.02-fold. It is likely that most of the U.S. incidence stems from sources unrelated to the gatherings, which could have also affected the counties with the gatherings. It is also possible that at least part of the U.S. incidence outside of gathering counties is due to COVID-19 spreading from the gathering counties.

Most counties saw an increase after the fifth day, which was shown in previous studies as a common time interval from COVID-19 infection to detection.^9^ The onset and peak of the incidence increase varied between counties, which could result from differences in testing speed and the effects of secondary infections.

The main limitation of our findings is that counties that held mass gatherings during the COVID-19 pandemic might be less likely to take other COVID-19 precautions that reduce COVID-19 transmission, such as masks, and keeping a distance of 6 feet.^10^ There have been reports of a lack of such precautions in the analyzed mass gatherings.^3^

Our findings suggest that mass gatherings may increase COVID-19 incidence even when they are held outdoors and in areas with low COVID-19 incidence. Therefore, the public health consequences of such mass gatherings should be considered. If such mass gatherings continue, preventive measures, such as masks, distancing, and contact tracing, should be taken in those gatherings to limit COVID-19 infections.

## Data sharing

We used publicly available data from the COVID-19 data repository of John Hopkins University, which is accessible at this link-https://github.com/CSSEGISandData/COVID-19. We are also prepared to share our data upon specific request to the corresponding author, Oren Miron.

The lead author (the manuscript’s guarantor) affirms that the manuscript is an honest, accurate, and transparent account of the study being reported; that no important aspects of the study have been omitted; and that any discrepancies from the study as planned (and, if relevant, registered) have been explained.

## Supporting information

STROBE checklist

## Data Availability

Data is publicly available at the John Hopkins University data repository on COVID-19

https://github.com/CSSEGISandData/COVID-19

## Author contribution

Concept and design: All authors.

Acquisition, analysis, or interpretation of data: All authors.

Drafting of the manuscript: OM.

Critical revision of the manuscript for important intellectual content: KHY, RWM, ND.

Statistical analysis of verified data: OM and KHY. Study supervision: ND.

OM is guarantor and corresponding author for this work, and attests that all listed authors meet authorship criteria and that no others meeting the criteria have been omitted.

## Ethic approval

Due to use of public, de-identified data, it is not required.

## Patient and public involvement

Patients or the public were not involved in the design, conduct, reporting, or dissemination plans of our research. All data was from de-identified existing public sources. We plan to share this with the public on social media.

